# Bantu on the brain: Community-informed adaptations of the Macarthur-Bates Communication Development Index to contemporary Zambian languages using item response theory

**DOI:** 10.1101/2025.06.03.25328935

**Authors:** Megan Genevieve Latoya, Peter C. Rockers, Carol Buumba, Frazer Shimaingwa, Donald M. Thea, Ethan Zulu, Julie M. Herlihy

## Abstract

The MacArthur-Bates Communication Development Inventories (MB-CDI) are a widely used set of tools to assess language acquisition in early childhood. Although it has been adapted in 120 languages, there is not a linguistic nor culturally congruent tool for any of the 72 Zambian Bantu languages. This manuscript describes the process to adapt the MB-CDI for two of the most spoken languages in Zambia: Nyanja and Bemba. Using mixed-methods with caregivers of children aged 16 to 30 months in Lusaka, Zambia, we have constructed two Bantu-language adaptations of the MD-CDI: Words and Sentences short form. Two focus group discussions of caregivers were conducted, one in Nyanja (n=10) and one in Bemba (n=10). The goal of the focus groups was to generate a list of commonly heard words in early childhood. Facilitators then administered individual assessment surveys to Bemba (n=77) and Nyanja (n=109) caregivers to determine which words from the generated lists their child (aged 16-30 months) had acquired in productive speech. We then fitted a 2-parameter item response theory model to our data to reflect 100 words, characterized by a range of difficulty and highest ability to effectively distinguish between different levels of language proficiency. Our adapted MB-CDI for Bemba and Nyanja is a language acquisition assessment tool for ages 16-30 months to measure language development in the two most spoken Bantu languages in Zambia.

## Introduction

Language acquisition is how a child learns to process speech, communicate needs, and adapt to their environment. It is a cornerstone of early literacy and foundational to social and cognitive neurodevelopment (1). Language acquisition is a dynamic process that involves genetics, environment, and culture. As global policy begins to prioritize early childhood development for vulnerable children, it is imperative to develop culturally and linguistically valid assessment tools which acknowledge the complex interplay between environment and biology.

The MacArthur-Bates Communicative Development Inventories (MB-CDI) are a widely-adapted set of parent-reported instruments to assess early language acquisition in young children (2). The suite of MB-CDI assessments are valid among children aged 8-30 months, including those who are developing atypically (3). It has been officially adapted 120 times, including for language groups found in Zambia’s neighbors Zimbabwe (4) and Malawi (5). The MB-CDI can accommodate low-literacy and low-resource settings because caregivers can be read the assessment aloud, the assessment can be completed by paper or online, and facilitators can be trained quickly. However, a major Eurocentric disparity in language coverage prevents widespread adoption— less than 1% of Bantu languages have been represented (6). As of this study, no adaptation exists for any of Zambia’s 72 national languages.

Appropriate adaptation of the MB-CDI steps beyond direct translation and strives for conceptual cohesion, aiming to find socioculturally appropriate items (7). Socioculturally appropriate vocabulary items refer to items and concepts that a child will have real experience with (so that a child is not expected to know the word spaghetti if it is not a food item they would encounter) and accurately reflect the nuances in language development in the child’s environment (such as “auntie” referring to all older females). As the capitol of Zambia, Lusaka is a notably multicultural city where hybridized “town” Nyanja (7) and “town” Bemba (8) have blended several languages and become widely adopted communication forms in everyday city life.

This manuscript describes the methods and outcome of adapting the short form of the MB-CDI: Words and Sentences (MB-CDI:WS) for the two most spoken Bantu languages in Zambia: Nyanja and Bemba, inclusive of town forms.

## Methods

### Study Design

This study was approved by BUSPH IRB (FWA00000338) and University of Zambia Biomedical Research Ethics Committee (NHRAR-REC No 2021-05-0002).

We conducted a two-phase mixed-method study (S1 Appendix) using focus group discussions and individual assessment surveys. We created the two proposed adaptations using the validated Chichewa language MB-CDI (5), due to sociocultural and linguistic similarities between Malawi and Zambia. To ensure conceptual preservation, each item in Chichewa was translated by a native speaker into our target local languages (Nyanja and Bemba), then back-translated into English.

Caregivers were recruited from the Maternal Child Health unit at Chawama First Level Hospital, a government health facility in Lusaka, Zambia. Inclusion criteria included: caregivers of a child aged 16-30 months, who primarily spoke either Nyanja or Bemba, and who were not graduates of the Zambian Infant Cohort Study previously described (9)(10). Recruitment began on January 1th, 2024 and lasted through February 12^th^, 2024. Participants provided written and oral consent at time of recruitment; participants who did not read or write provided consent via thumb print in the presence of a witness.

Focus groups generated inventories of words that children would be commonly exposed to in early childhood. Facilitators read each vocabulary item aloud from the translated Chichewa inventories and asked caregivers to indicate by hand if their child would hear each word in daily life. Caregivers also provided alternative context-specific suggestions for each item. Facilitators then asked the caregivers to generate words their children have heard in accordance with 12 predefined categories (e.g. common household items, animals). We derived a list of 135 (Bemba) and 131 (Nyanja) potential vocabulary items for inclusion on the individual assessment inventory, representing 126 Bemba and 120 Nyanja unique items after condensing for regional spelling variation, town, and urban dialects.

In the second phase of the study, we conducted individual assessment surveys to test productive speech ability-i.e. if a child could *independently say* each suggested vocabulary item. Staff read each item aloud to the caregiver and asked if their child has independently used the vocabulary item. Every 25^th^ word, staff also elicited the context in which a child said the word to confirm independent usage. To account for the multi-lingual environment of urban Zambia, children were credited with acquisition of the word when parental report confirms their use in any language. To account for ongoing development of the soft palate, children were similarly credited if they attempted to say a word but could not correctly pronounce it; or, if they used a ‘baby version’ (i.e. ‘dada’ for ‘daddy’). Children were not credited if they never independently used the vocabulary item, even if they could repeat it. All data were collected in REDCap, a password-protected and encrypted software.

### Statistical analysis

Data were analyzed using STATA version 18 (11). Demographic data were collected and stratified according to primary language spoken.

We used item response theory (IRT) to analyze individual assessment data. IRT has been used to create short adapted MB-CDI forms with high validity (12) (13). We fitted a 2-parameter IRT model to describe key parameters of each vocabulary item: difficulty and discrimination (14). In our study difficulty describes the challenge an item presents for children at different developmental stages, while discrimination indicates how well an item can distinguish between two children with varying levels of language ability. See S2 Appendix for analysis design schematic.

We then graphically examined the logistic function of each item’s information. From these models, we selected 100 words in each language which 1) taken together, accurately capture a wide range of difficulty and 2) have the highest discrimination values. We then modeled the 100-word dataset. The final 100-word models served as the adapted MB-CDI:WS short forms in Nyanja and Bemba, respectively.

## Results

### Focus Group Discussion

We conducted one focus group in Nyanja and one in Bemba. Each focus group discussion had 10 participants. We elicited a total of 131 Nyanja and 136 Bemba vocabulary items. Several items suggested were the “deep” and “town” forms of a single suggested item, reflecting urban hybridity. Items which referred to the same concept in both forms of the language were collapsed to create 120 Nyanja and 126 Bemba words on the inventory, respectively.

### Individual Assessment

A total of 77 Bemba caregivers and 123 Nyanja caregivers completed the assessment. Three primarily-speaking Nyanja children were excluded due to missing birthdate. Ten children in the Nyanja dataset were excluded due to falling out of the age parameters (16 to 30 months). One child in the Bemba dataset was excluded due to individual assessment missing data. The final datasets were composed of 76 Bemba speakers and 109 Nyanja speakers, respectively.

Caregivers had the option of responding “Yes”, “No”, and “I don’t know” to each vocabulary item in the individual assessment.

Distributions of baseline demographic characteristics by primary language spoken are displayed below:

**Table 1:** Demographic Data for BOBS Individual Assessment Participants.

| Characteristic | Nyanja (N= 110) | Bemba (N=76) |
| --- | --- | --- |
|  | *Mean, STE* | *Mean, STE* |
| Caregiver Age (years) | 28.2 (7.5) | 28.4 (6.6) |
| Child’s Age (months) | 23.5 (4.3) | 22.6 (4.2) |
|  | **% (N)** | **% (N)** |
| Caregiver Gender (female) | 100 (110) | 100 (76) |
| Child’s Gender (female) | 54.5 (60) | 55.3 (42) |
| Caregiver Relationship to the Child |  |  |
| *Parent* | 100 (110) | 98.7 (75) |
| *Grandparent* | 0 (0) | 1.3 (1) |
| *Other* | 0 (0) | 0 (0) |
| Household Language Exposure |  |  |
| *Unilingual* | 87.3 (96) | 78.9 (60) |
| *Bilingual* | 12.7 (14) | 17.2 (13) |
| *Trilingual* | 0.0 (0) | 3.9 (3) |

The average age of Nyanja- and Bemba-speaking children were 23.5 and 22.6 months, respectively. Most caregivers were the parents of the children (100% Nyanja, 98.7% Bemba); grandparents were the other family relationship represented (0% Nyanja, 1.3% Bemba). Most (87.3% Nyanja, 78.9% Bemba) of the caregivers the study stated that their children were exposed to one language. 12.7% of Nyanja-speaking caregivers stated that more than one language was spoken in the household-the most common secondary language for primarily Nyanja-speaking children was English (8 out of 14 children with bilingual exposure), followed by Bemba (6 out of 14 children with bilingual exposure). 17.2% of Bemba-speaking children were exposed to a second language regularly; the most commonly reported second language was Nyanja (13 of 14 children with bilingual exposure) followed by English (1 of 14). One Bemba-speaking caregiver reported that their child was exposed to a second language other than Nyanja or English but did not specify. Three (3.9%) primarily Bemba-speaking caregivers reported trilingual exposure in Bemba, Nyanja, and English.

### IRT

Nyanja: The first 2PL IRT model with the entire dataset did not reach convergence; inspecting the data showed two items with non-variance (100% “yes”) and two items with very little variance in responses. Items with little to no variance are “too easy” or “too hard” for children 16-30 months to meaningfully contribute to the model. We achieved convergence after dropping two items from the Nyanja dataset. From this model we chose 100 words with the highest discrimination values and modeled the final lists. The finalized 2PL models of our 100-word lists is provided in Appendix 3.1. Discrimination and difficulty values for each IRT iteration are available in Appendix 2.1.

Bemba: The first 126-item model also did not reach convergence due to three items with non-variance. Convergence was achieved after we dropped five total items (Appendix 3.1); we again modeled 100 words with the highest discrimination value. Discrimination and difficulty values for each IRT iteration are available in Appendix 3.1. The finalized 2PL models of our 100-word list is provided (Appendix 3.2).

Graphic modeling: We generated item characteristic curves (ICCs) and item information functions (IIFs) of the final 100-word lists. ICCs (Figs 1 and 2) display the spread of difficulty the list creates; IIFs (Figs 3 and 4) show the amount of “information” each item contributes to the model at a given language ability.

**Fig 1:**
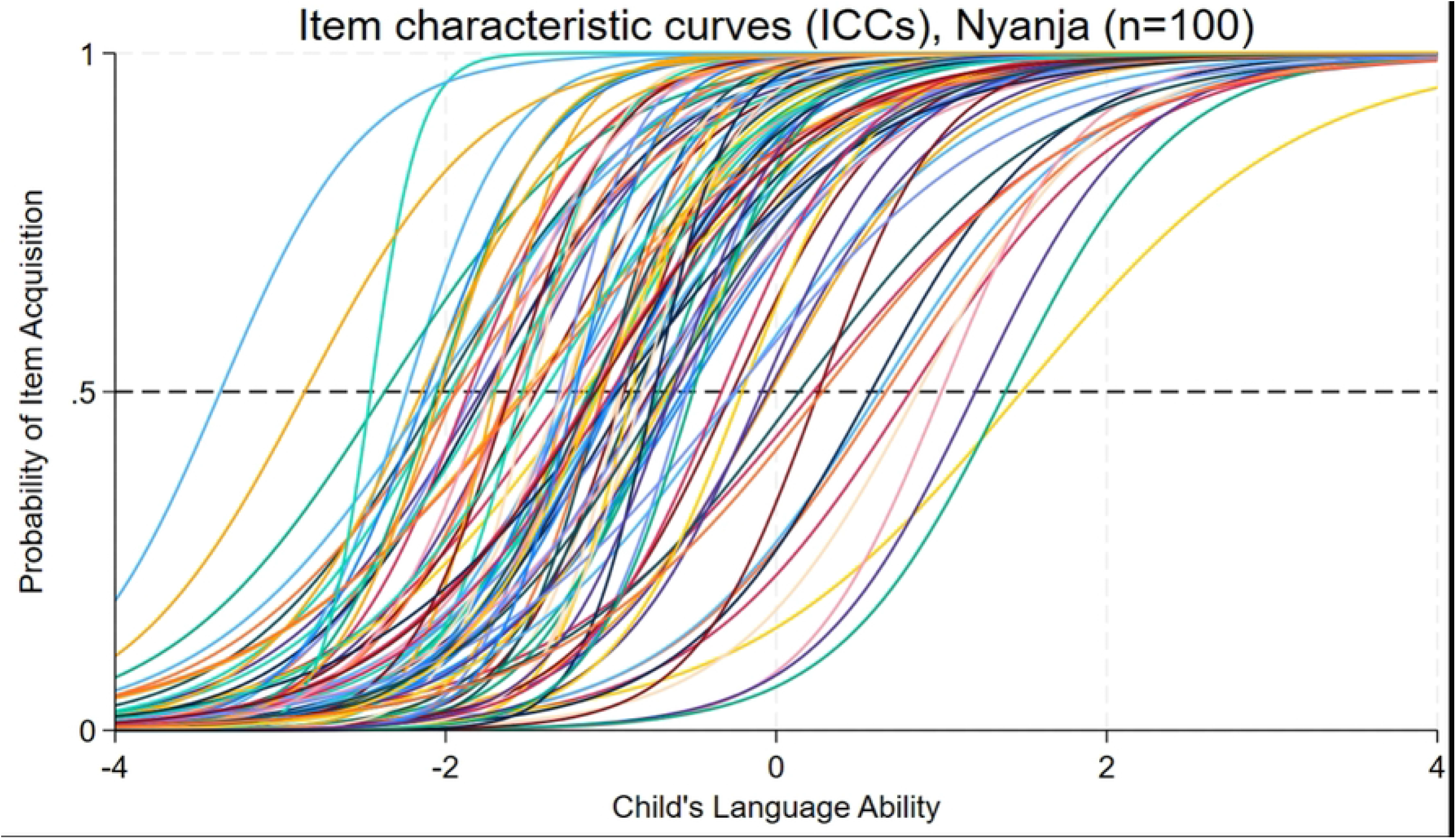
Item Characteristic Curves of final 100 words in Nyanja

**Fig 2:**
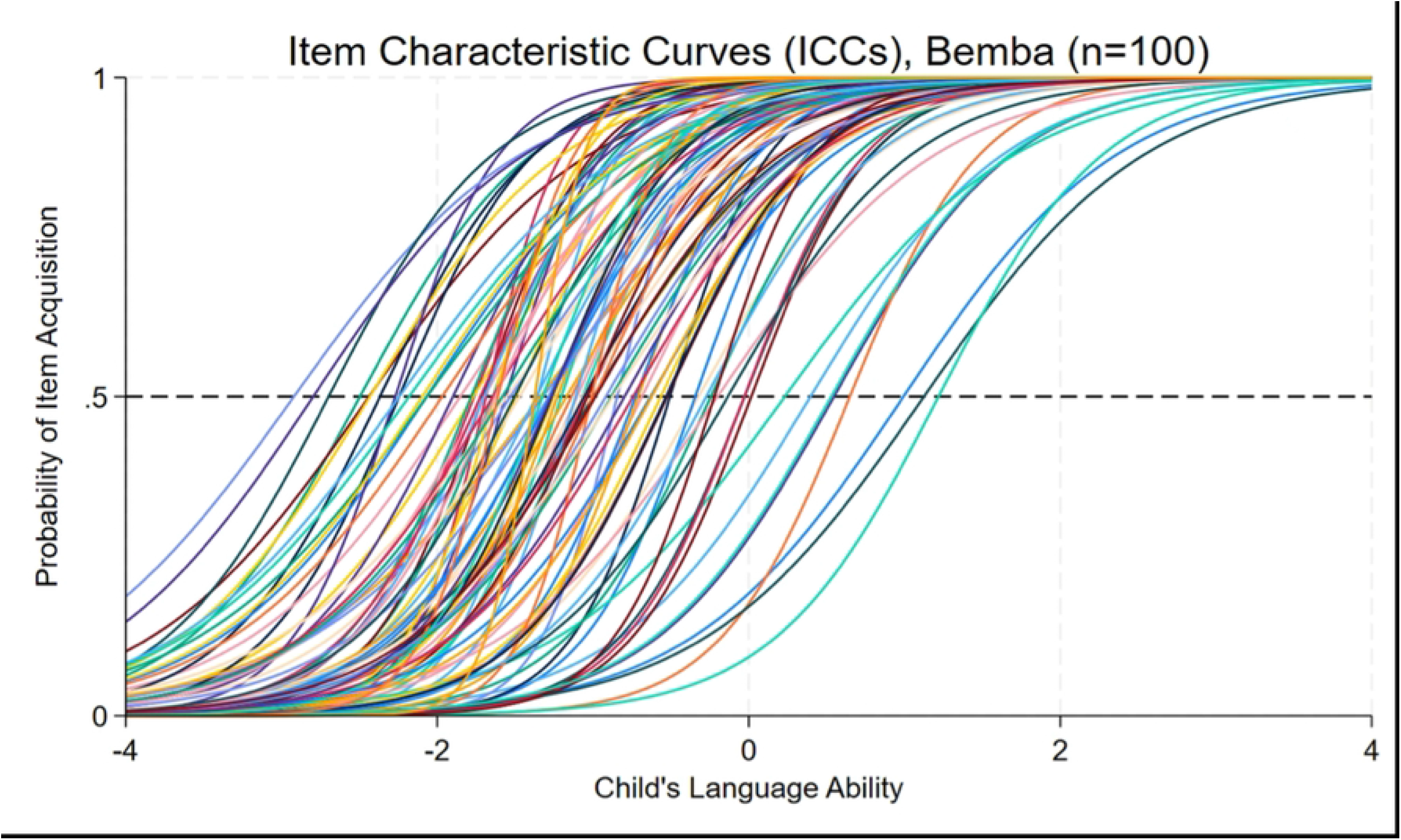
Item Characteristic Curves of final 100 words in Bemba

**Fig 3:**
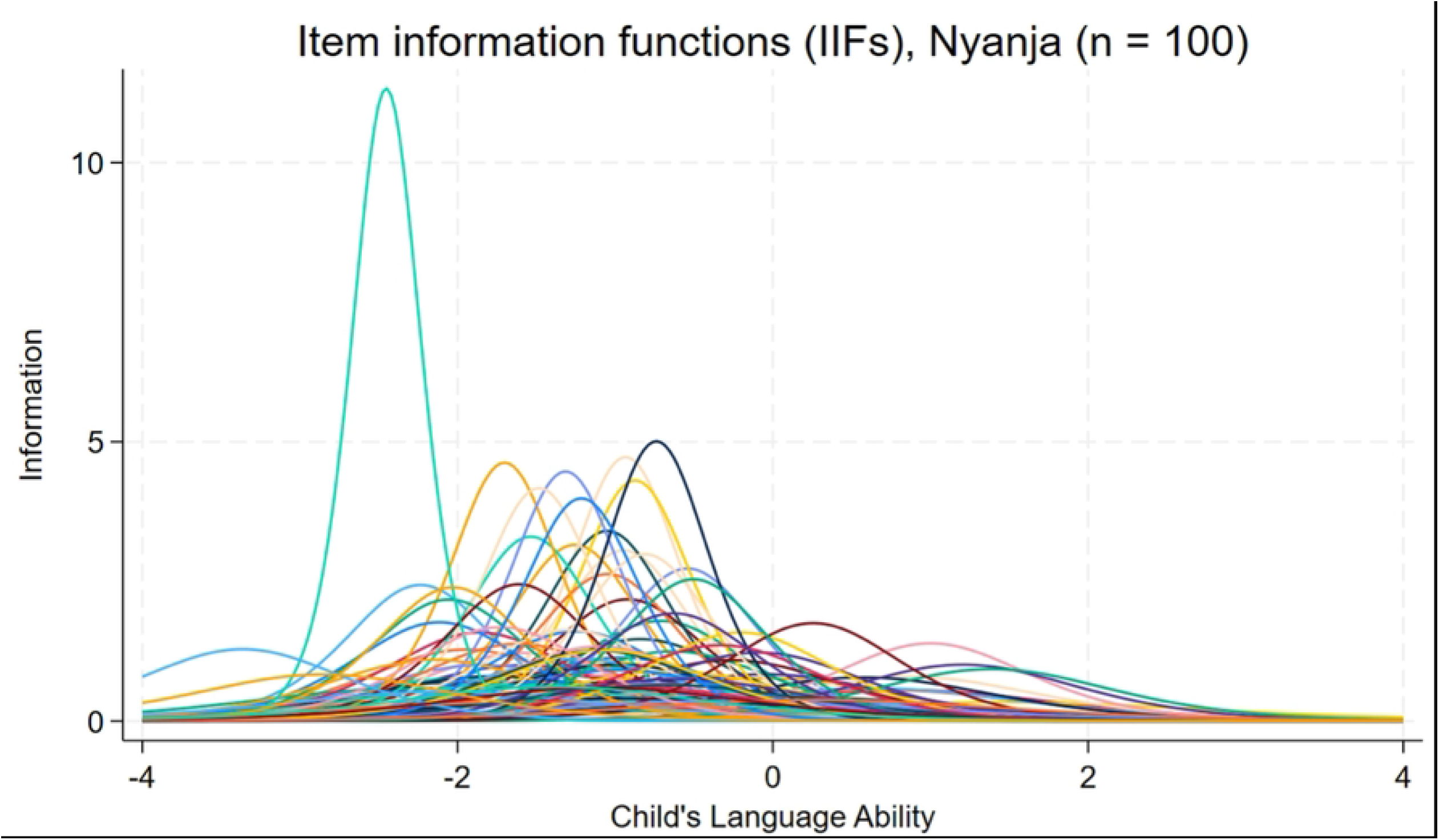
Item information Functions of final 100 words in Nyanja

**Fig 4:**
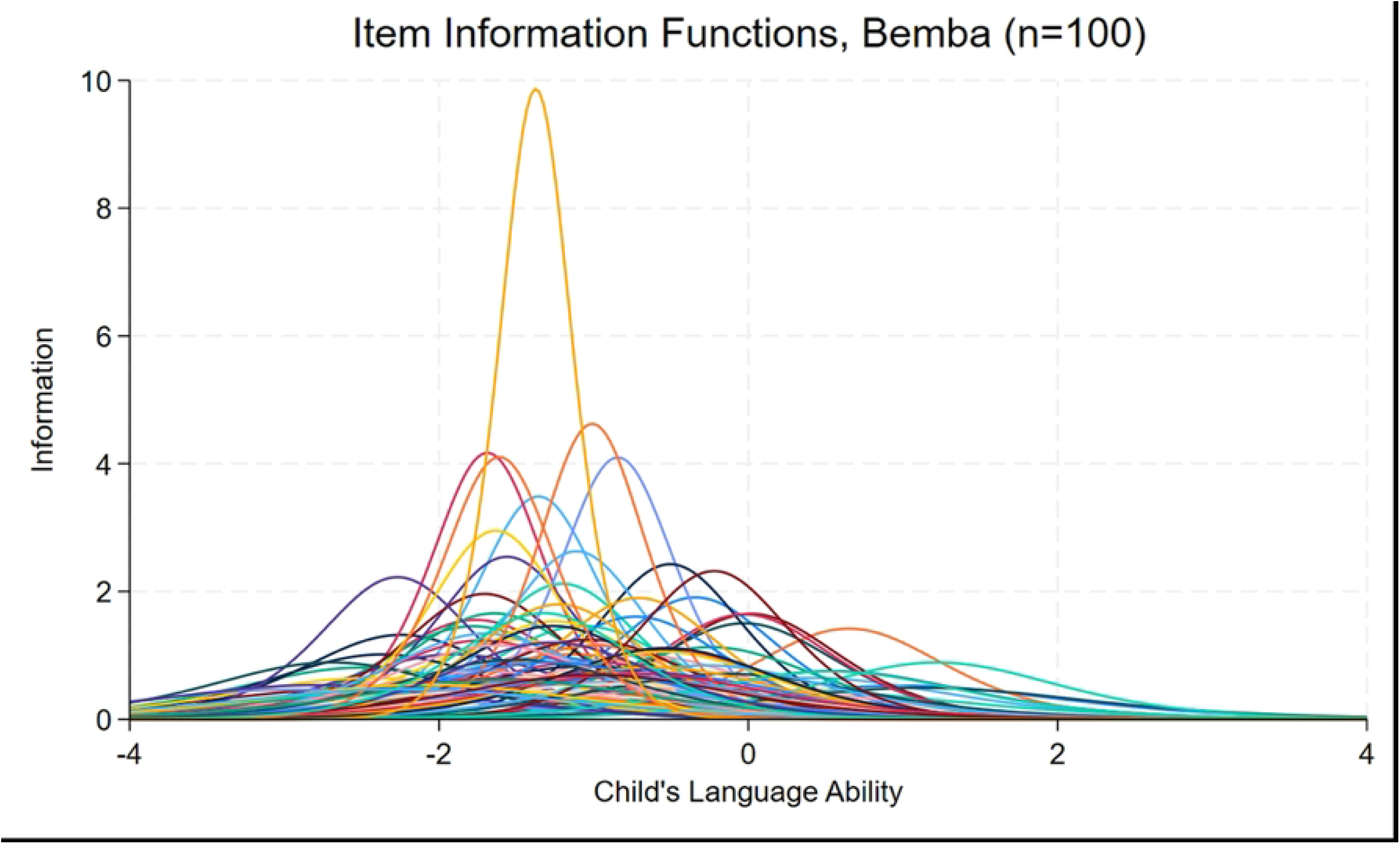
Item Information Functions of final 100 words in Bemba

ICC graphs generated by IRT analysis show item difficulty as the position of the curve along ability (x-axis) at which a child has a 50% chance of acquiring an item (reference line on y-axis), while the steepness of the curve indicates an item’s discrimination at a given difficulty level (15).Graphs of IIFs (Figs 3 and 4) show the level of information an item contributes to determining a person’s ability at a given level, while peaks of the IIF show an item’s discrimination value (Rosider, n.d.) The IIF curves with the highest peaks indicate items most effective at discriminating between children around the item’s difficulty level.

## Discussion

In this study we combined a community-informed approach and item response theory analysis to create a precise, sociolinguistically congruent adaptation of the MB-CDI:WS short form for the two most spoken Bantu languages in Zambia. Our model accommodates for both urbanized “town” forms and “deep” traditional linguistic structures present in diverse Zambian locales. Item response theory informed our choice of vocabulary item selection to present a variety of difficulty levels with the best ability to discriminate between children’s differential neurodevelopmental progress. The resulting tailored inventory is responsive to the nuances of early language development in Bantu-speaking environments. Both forms have been accepted as official adaptations by the MB-CDI Advisory Board.

Until this study, no language-specific neurodevelopmental assessment has been created to measure the acquisition of Zambian languages. As Zambian national strategy has recently begun to prioritize early childhood development as a major goal (16), it is imperative that a suite of culturally congruent tools can accurately measure a child’s progress within their peer group.

Study limitations include use of a convenience sample within one compound of Lusaka; however, a randomized sample was beyond the scope of this study and the peri-urban compounds of Lusaka have large numbers of Bamba and Nyanja speakers. We also used IRT analysis to choose words with the highest precision. In the IRT estimation process, the calibrating group’s influence on the scale is minimized by adjusting for the mean and spread of ability within the sample (15).

Additionally, our samples were underpowered. The IIFs generated from our final 100-word models still show low information peaks for several items. A larger sample may increase the information each item contributes to the model. And while our adaptation is currently being piloted in the Zambia Infant Cohort Study to measure neurological outcomes of children who have been exposed to HIV, it still needs further norming to maximize its usefulness in cross-cultural neurodevelopmental research. Further suggestions also include validation of the adapted protocol against accepted neurodevelopmental assessments and integration into the MB-CDI Wordbank open database.

Despite these limitations, our study has shown that mixed-methods approach and statistical modelling can be used to elicit a precise list of socioculturally appropriate items for underrepresented languages globally. Our use of focus groups generated several alternative items quickly, as well as gave space to understand the reasoning for item preference. Our use of IRT to select the final items from a pool of community-based suggestions created a short, tailored tool.

Our MB-CDI:WS short form adaptation can be used by other researchers to compare the neurodevelopment of different populations with instruments that measure children against their peers. The finalized instrument allows for assessment of children’s language acquisition for Nyanja and Bemba speaking children in a variety of contexts. Sociolinguistically valid tools can capture the dynamic flexibility of contemporary African cities and help to inform early childhood development research globally.

## Data Availability

Availability of data and materials: Data were analyzed in STATA 18. Materials and analysis code are available at Harvard Dataverse https://doi.org/10.7910/DVN/R26ULF. This study is not preregistered.

## Acknowledgments

We would like to acknowledge to hard work and dedication of our data collection team: Gift Hope, Justina Zulu, Esnala Chibanda, Priscilla Katiba, Namenda Nasilele, Dorcas Mulenga, Ba Lyness Kamanga, Vivian Fulilwa, Namwiya Jaemmata, and Charles Nthele. We would also like to thank Bizu Gelaye, PhD, MPH, for his invaluable expertise on item response theory applications. We are of course incredibly grateful for the steadfast guidance of Phillip Dale, PhD throughout the process of developing our adaptation. Most importantly, we are forever indebted to the women and children who shared their lives with our team; thank you.

---

**S1 Appendix: Study design schematic**

**S2 Appendix: Study Analysis Schematic**

**S3 Dataset: 2-parameter IRT model, Nyanja**

**S4 Dataset: 2-parameter IRT model, Bemba**

**S5 Appendix: MB-CDI: WS short-form, Nyanja**

**S6 Appendix: MB-CDI: WS short-form, Bemb**

## Notes

### Competing Interest Statement

The authors have declared no competing interest.

